# Reliability of single-lead electrocardiogram interpretation to detect atrial fibrillation: insights from the SAFER Feasibility Study

**DOI:** 10.1101/2024.01.29.24301927

**Authors:** Katie Hibbitt, James Brimicombe, Martin R. Cowie, Andrew Dymond, Ben Freedman, Simon J Griffin, FD Richard Hobbs, Hannah Clair Lindén, Gregory Y. H. Lip, Jonathan Mant, Richard J. McManus, Madhumitha Pandiaraja, Kate Williams, Peter H. Charlton, the SAFER Investigators

**Affiliations:** Department of Public Health and Primary Care, University of Cambridge, Cambridge, CB1 8RN, UK; Royal Brompton Hospital (Guy’s and St Thomas’ NHS Foundation Trust), Sydney Street, London, SW3 6NP, UK; Zenicor Medical Systems AB, 113 59 Stockholm, Sweden; Liverpool Centre for Cardiovascular Science at University of Liverpool, Liverpool John Moores University and Liverpool Heart & Chest Hospital, Liverpool, United Kingdom; and Danish Center Health for Services Research, Department of Clinical Medicine, Aalborg University, Aalborg, Denmark; Heart Research Institute, University of Sydney, Sydney 2006, Australia; Nuffield Department of Primary Care Health Sciences, University of Oxford, Oxford, OX2 6GG, UK

**Keywords:** atrial fibrillation, diagnosis, electrocardiogram, inter-rater agreement, screening

## Abstract

**Background and Aims:** Single-lead electrocardiograms (ECGs) can be recorded using widely available devices such as smartwatches and handheld ECG recorders. Such devices have been approved for atrial fibrillation (AF) detection. However, little evidence exists on the reliability of single-lead ECG interpretation. We aimed to assess the level of agreement on detection of AF by independent cardiologists interpreting single lead ECGs, and to identify factors influencing agreement.

**Methods:** In a population-based AF screening study, adults aged ≥65 years old recorded four single-lead ECGs per day for 1-4 weeks using a handheld ECG recorder. ECGs showing signs of possible AF were identified by a nurse, aided by an automated algorithm. These were reviewed by two independent cardiologists who assigned participant- and ECG-level diagnoses. Inter-rater reliability of AF diagnosis was calculated using linear weighted Cohen’s kappa (*κ_w_*).

**Results:** Out of 2,141 participants and 162,515 ECGs, only 1,843 ECGs from 185 participants were reviewed by both cardiologists. Agreement was moderate: *κ_w_* = 0.42 (95% CI, 0.32 – 0.52) at participant-level; and *κ_w_* = 0.51 (0.46 – 0.56) at ECG-level. At participant-level, agreement was associated with the number of adequate-quality ECGs recorded, with higher agreement in participants who recorded at least 67 adequate-quality ECGs. At ECG-level, agreement was associated with ECG quality and whether ECGs exhibited algorithm-identified possible AF.

**Conclusion:** Inter-rater reliability of AF diagnosis from single-lead ECGs was found to be moderate in older adults. Strategies to improve reliability might include participant and cardiologist training and designing AF detection programmes to obtain sufficient ECGs for reliable diagnoses.

**What’s New:**

- We observed moderate agreement between cardiologists when diagnosing AF from single-lead ECGs in an AF screening study.
- This study indicates that for every 100 screening participants diagnosed with AF by two cardiologists, there would be complete disagreement over the diagnosis of 70 further participants.
- We found that the quality of ECG signals greatly influenced the reliability of single-lead ECG interpretation.
- In addition, when multiple ECGs were acquired from an individual, the reliability of participant-level diagnoses was influenced by the number of adequate-quality ECGs available for interpretation.

## 1. Introduction

The electrocardiogram (ECG) is a fundamental technique for assessing the functionality of the heart. The process for recording a 12-lead ECG was described 70 years ago (1), and to this day the 12-lead ECG remains widely used for the diagnosis and management of a range of heart conditions (2). Whilst the 12-lead ECG is highly informative, providing several ‘views’ of the heart’s electrical activity, it can only be measured by clinicians in a healthcare setting. Recently, clinical and consumer devices have become available which allow individuals to record a single-lead ECG on demand via a smartwatch or handheld device (29). This approach has a number of useful features: such ECGs can be measured by patients themselves with no clinical input, can be acquired synchronously with symptoms, can be repeated on multiple occasions with minimal inconvenience and can be transmitted electronically to healthcare providers (3).

Atrial fibrillation (AF) is a common arrhythmia which confers a fivefold increase in the risk of stroke (4) which can be mitigated through anticoagulation (5). A significant proportion of AF remains unrecognised (6) as it may be asymptomatic or occur only intermittently. Self-captured, single-lead ECGs could greatly assist in the detection of AF (7) when: (i) used by device owners, with ECGs acquired opportunistically, upon symptoms, or when prompted by a device (8); and when (ii) used in screening programmes, allowing multiple ECGs to be acquired from an individual over a period of weeks (7). Indeed, European Society of Cardiology guidelines support the use of single-lead ECGs acquired from wearable or mobile devices to identify AF (8). Whilst automated algorithms can be used to identify those ECGs which show evidence of AF and therefore warrant clinical review (9), a final diagnosis of AF must be made by a physician interpreting an ECG (8). To date, there is little evidence on the reliability of single-lead ECG interpretation for AF diagnosis, and most existing evidence is derived from ECGs collected from hospital patients (10–13).

We aimed to assess the level of agreement on detection of AF by independent cardiologists interpreting single lead ECGs, and to identify factors which influence agreement.

## 2. Methods

We assessed inter-rater agreement using ECG data collected in a population-based AF screening study, in which each participant recorded multiple ECGs. Agreement between cardiologist interpretations was assessed at the participant-level (*i.e.* the overall participant diagnosis) and the ECG-level (*i.e.* interpretations of individual ECGs). In addition, we investigated the influence of several factors on the level of agreement (*e.g.* participant age and ECG quality).

### 2.1 Data collection

We collected the data for these analyses in the SAFER (Screening for Atrial Fibrillation with ECG to Reduce stroke) Feasibility Study (ISRCTN 16939438), conducted in 2019 and approved by the London-Central Research Ethics Committee (REC ref: 18/LO/2066). Participants were older adults aged ≥ 65 years old, who were not receiving long-term anticoagulation for stroke prevention, not on the palliative care register, and not resident in a nursing home. All participants gave written informed consent.

In this study, older adults (aged 65 and over) recorded single-lead ECGs at home using the handheld Zenicor EKG-2 device (Zenicor Medical Systems AB) (9). This device measures a 30-second, single-lead ECG between the thumbs, using dry electrodes. Participants were invited to attend a screening visit at their general practice, where a practice nurse showed the participant how to use the device, and supervised the participant recording the first ECG including reviewing ECG quality (28). Participants were then asked to record four ECGs per day for either 1, 2 or 4 weeks. The ECGs were transferred to a central database for analysis and review.

Participant- and ECG-level diagnoses were obtained as follows (and as summarised in Figure 1). First, a computer algorithm was used to identify abnormal ECGs (Cardiolund ECG Parser algorithm, Cardiolund AB, Sweden). The algorithm has previously been found to have a sensitivity for AF detection of approximately 98% (9). Second, a nurse reviewed all the ECGs which were classified by the algorithm as abnormal, and manually corrected any algorithm misclassifications based on their clinical judgement. The nurse then identified participants for cardiologist review as those participants with at least one ECG classified as abnormal which the nurse deemed exhibited signs of possible AF (as detailed in (14)). Third, these participants were sent for review by two highly experienced cardiologists, both of whom had substantial ECG reviewing experience including reviewing single-lead ECGs acquired by handheld devices using dry electrodes (GYHL and MRC). The cardiologists had access to all the ECGs from these participants, though it was not anticipated that ECGs that were classified as normal would be reviewed, or that all abnormal ECGs would be reviewed, once a participant-level diagnosis had been reached. Each cardiologist independently provided a diagnosis for each participant. For AF to be diagnosed it was required to be present for the whole 30 seconds, or the entire trace where the ECG was interpretable. No other formal definition of AF was provided for the cardiologists to use. In addition, on an *ad hoc* basis, the cardiologists also provided diagnoses for individual ECGs and labelled ECGs as ‘low-quality’. Diagnoses were categorised as: AF ≥30 seconds duration; cannot exclude AF; or, non-AF. Labels of ‘low-quality’ were typically provided where there was baseline wander or artifact making the rhythm uninterpretable by the cardiologist.

**Figure 1:**
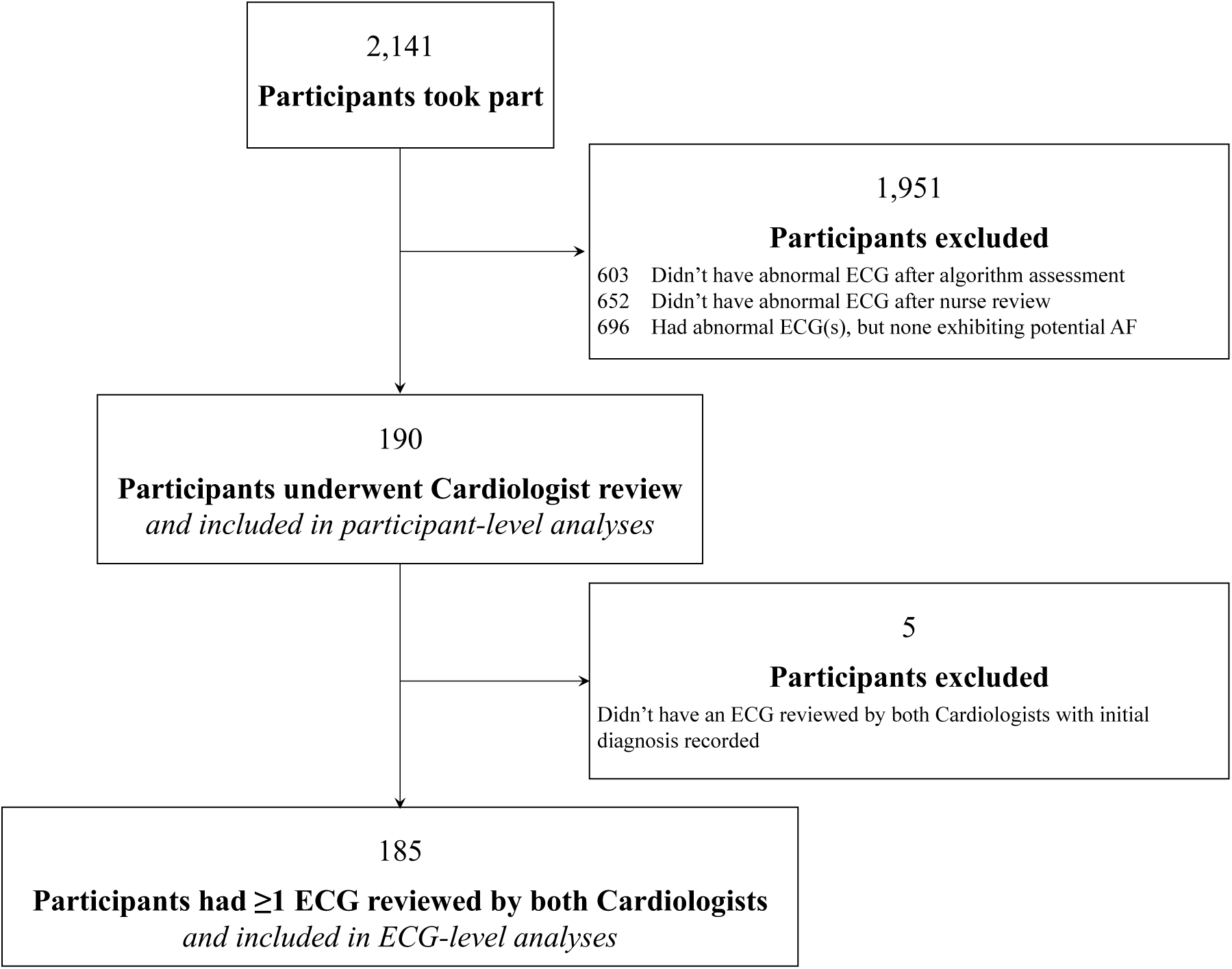
Data selection at the participant-level.

We extracted a subset of the collected data for the analysis as follows. Only data from those participants who were reviewed by both cardiologists were included in participant-level analyses. In addition, only those ECGs which were reviewed by both cardiologists were included in ECG-level analyses. We excluded from analyses any ECGs for which a cardiologist’s initial diagnosis was not recorded (prior to subsequent resolution of disagreements).

### 2.2 Data Processing

We obtained the characteristics of each ECG as follows: First, the computer algorithm extracted the following characteristics: heart rate, ECG quality (either normal or poor quality), level of RR-interval variability (calculated as the standard deviation of RR-intervals divided by the mean RR-interval), and whether or not an ECG exhibited algorithm-identified possible AF (defined as the ECG having either irregular RR-intervals or a fast regular heart rate). Second, the quality of ECGs was obtained by combining the quality assessment provided by the algorithm with cardiologist comments on ECG quality: any ECGs which the algorithm or at least one cardiologist deemed to be of poor quality were classed as low quality in the analysis. ECGs for which the algorithm was unable to calculate heart rate or RR-interval variability were excluded from analyses requiring those characteristics.

### 2.3 Statistical Analysis

We assessed the reliability of ECG interpretation using both participant-level diagnoses and ECG-level cardiologist diagnoses. First, we reported the overall levels of agreement. Second, we assessed the influence of different factors on levels of agreement, such as the influence of ECG quality. The factors assessed at the participant-level were: age, gender, number of adequate-quality ECGs recorded by a participant, and the number of ECGs recorded by a participant exhibiting algorithm-identified possible AF. The factors assessed at the ECG-level were: heart rate, RR-interval variability, ECG quality, and whether or not an ECG exhibited algorithm-identified possible AF. We investigated factors which were continuous variables (such as heart rate) by grouping values into categories with similar sample sizes (*e.g.* heart rates were categorised as 30-59 bpm, 60-69 bpm, etc).

We assessed agreement between cardiologists using inter-rater reliability statistics. The primary statistic, Cohen’s kappa, *κ*, provides a measure of the difference between the actual level of agreement between cardiologists, and the level of agreement that would be expected by random chance alone. Values for *κ* range from -1 to 1, with -1 indicating complete disagreement; 0 the level expected by chance; 0.01-0.20 slight agreement; 0.21-0.40 fair agreement; 0.41-0.60 moderate agreement; 0.61-0.80 substantial agreement; 0.81-0.99 almost perfect agreement; and 1 perfect agreement (15). The second statistic, a weighted Cohen’s kappa, *κ_w_*, reflects the greater consequences of a disagreement of ‘AF’ vs ‘non-AF’, compared to a disagreement of ‘cannot exclude AF’ vs either ‘AF’ or ‘non-AF’. We weighted disagreements of ‘AF’ vs ‘non-AF’ as complete disagreements, whereas disagreements including ‘cannot exclude AF’ were weighted equivalently to the level expected by chance. We reported the third statistic, percentage agreement, to facilitate comparisons with previous studies.

We calculated 95% confidence intervals for *κ* and *κ_w_* using bootstrapping. We undertook tests for significant associations between factors (*e.g.* heart rate) and the level of agreement using a chi-square test for independence between the proportion of agreement in each category.

## 3. Results

2,141 participants were screened, who recorded a total of 162,515 ECGs. Of the 2,141 participants who were screened, 190 had ECGs which underwent cardiologist review and were therefore included in the participant-level analyses, as shown in Figure 1. Most participants’ ECGs were not sent for cardiologist review (1,951 participants) because either: (i) the computer algorithm did not find any abnormalities in their ECGs (603 participants); or (ii) the nurse reviewer judged that none of their abnormal ECGs exhibited signs of possible AF (1,348 participants).

The 190 participants whose ECGs underwent cardiologist review recorded a total of 15,258 ECGs, with a median (lower – upper quartiles) of 67.0 (56.0 - 112.0) ECGs each. The two cardiologists assigned diagnoses to 1,996 and 4,411 of these ECGs respectively of which 1,872 ECGs were assigned diagnoses by both cardiologists. Initial diagnoses (prior to subsequent resolution of disagreements) were not recorded for 29 of these ECGs, leaving 1,843 available for ECG-level analyses (see Figure 2). The difference in the number of ECGs assigned diagnoses by each cardiologist demonstrates their different approaches to reviewing, with the cardiologists assigning diagnoses to 9.8 (3.9-18.5) % vs. 18.6 (6.8-48.8) % of participants’ ECGs.

**Figure 2:**
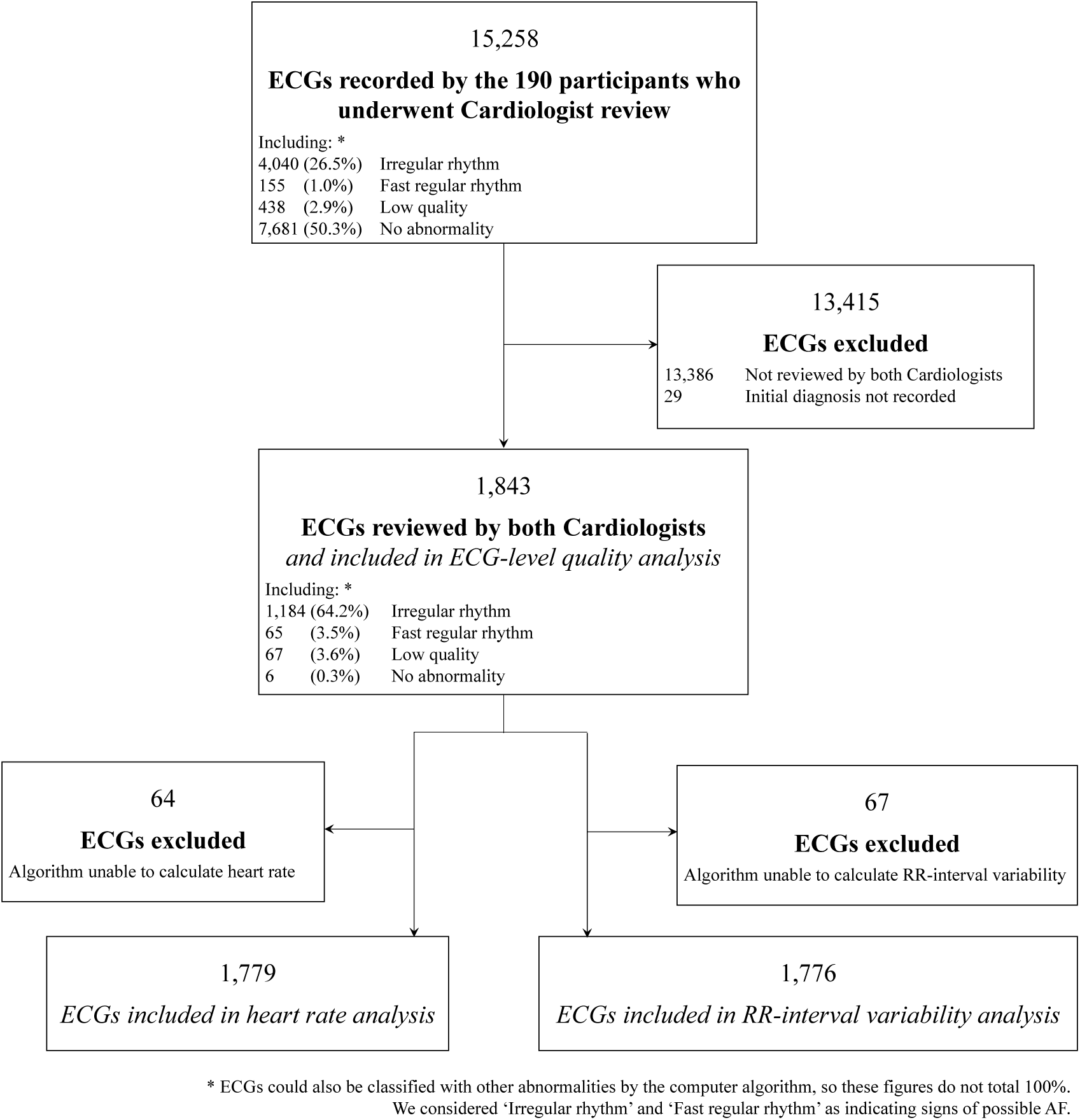
Data selection at the ECG-level.

### 3.1. Reliability of AF diagnosis at the participant-level

The inter-rater reliability of AF diagnosis at the participant-level, when the cardiologists had access to all the ECGs recorded by a participant, was moderate (*κ_w_* = 0.48 (0.37 − 0.58); *κ* = 0.42 (0.32 − 0.52); and %*_agree_* = 66.3% (Table 1).

**Table 1:**
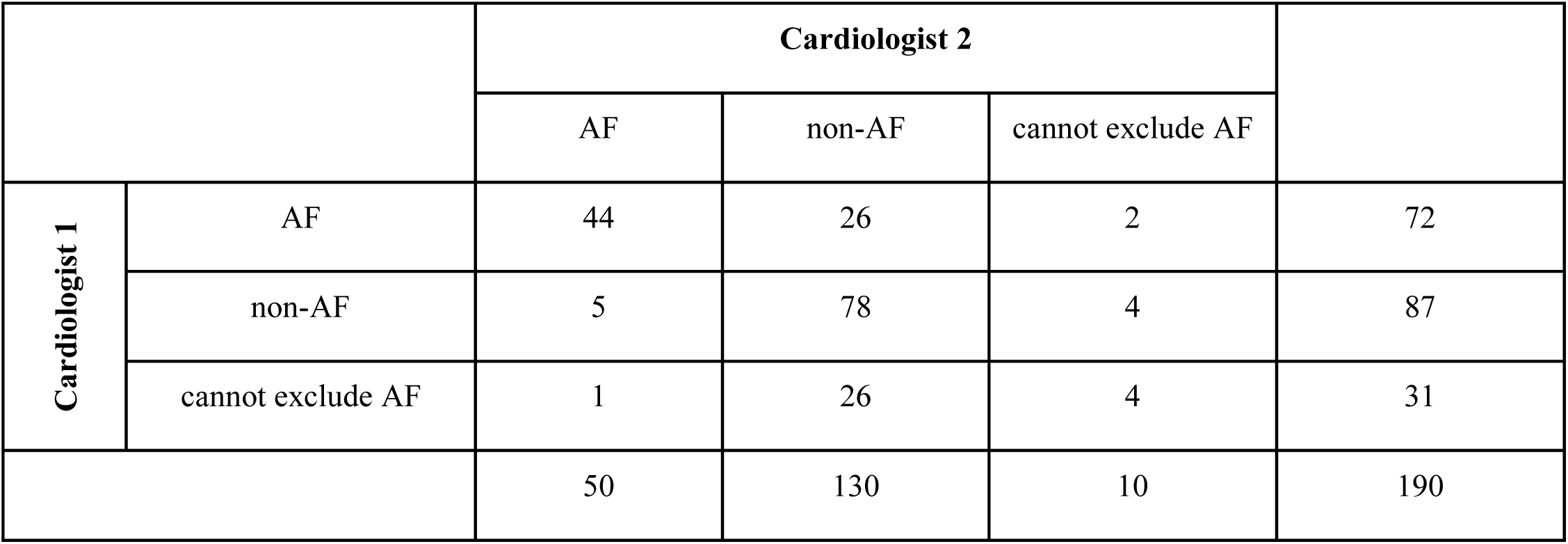
Agreement between cardiologists on participant-level AF diagnoses.

|  |  | Cardiologist 2 |  |  |  |
| --- | --- | --- | --- | --- | --- |
|  |  | AF | non-AF | cannot exclude AF |  |
| Cardiologist 1 | AF | 44 | 26 | 2 | 72 |
|  | non-AF | 5 | 78 | 4 | 87 |
|  | cannot exclude AF | 1 | 26 | 4 | 31 |
|  |  | 50 | 130 | 10 | 190 |

The results for the relationship between the level of agreement between cardiologists and factors at the participant- and ECG-level are presented in Figure 3 and Table 2. At the participant-level, the level of agreement was significantly associated with the number of adequate-quality ECGs recorded by a participant. Participants who recorded at least 67 adequate-quality ECGs had a significantly higher level of agreement in their diagnoses than those who recorded fewer than 67. There was agreement on 52.6% of participant-level diagnoses in those participants with <67 adequate-quality ECGs, compared to 80.0% in those with 67 or more. Of the 31 participants for whom there was complete disagreement (where one cardiologist diagnosed AF and the other diagnosed non-AF), 23 (74%) recorded <67 adequate-quality ECGs. There was no significant association between the level of agreement and age or gender.

**Figure 3:**
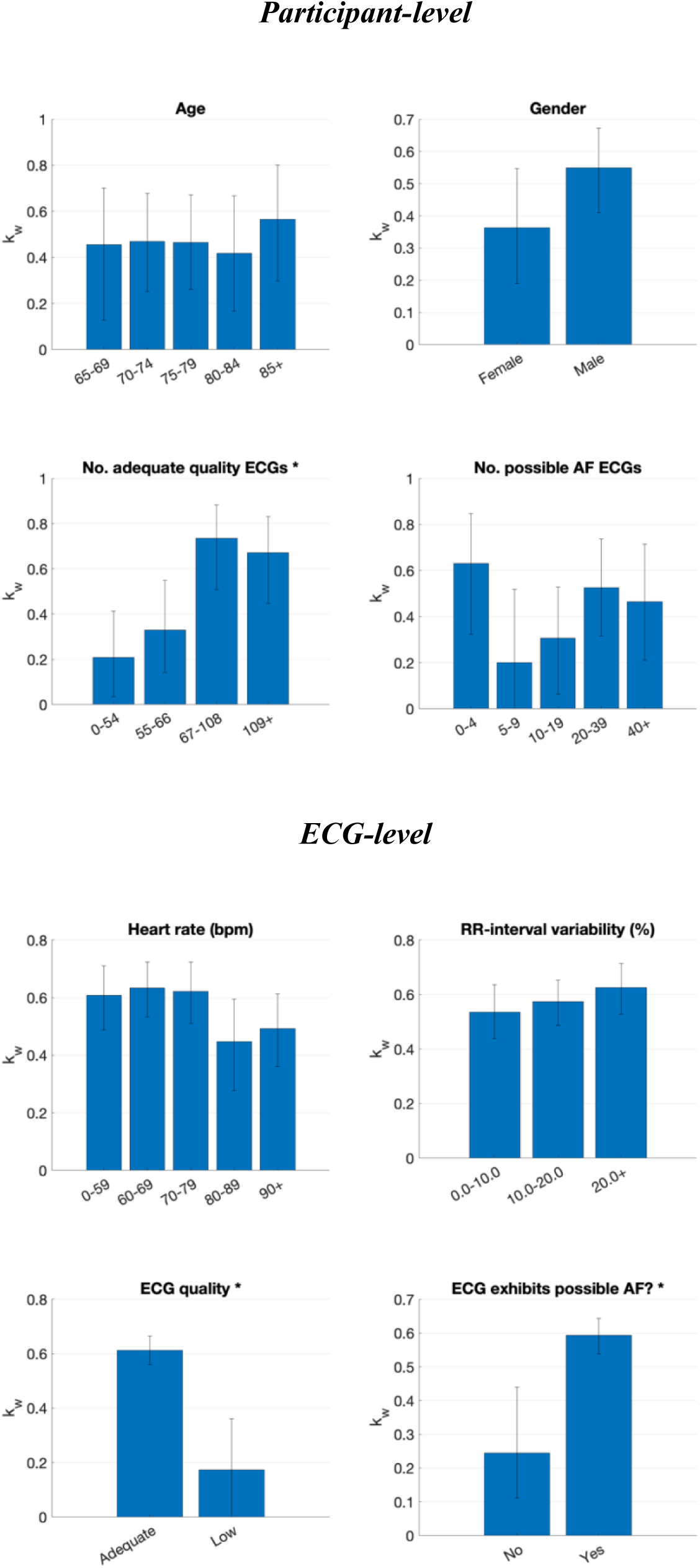
Relationships between the level of agreement between cardiologists and factors at the participant- and ECG-level. (* denotes a significant association)

**Table 2:**
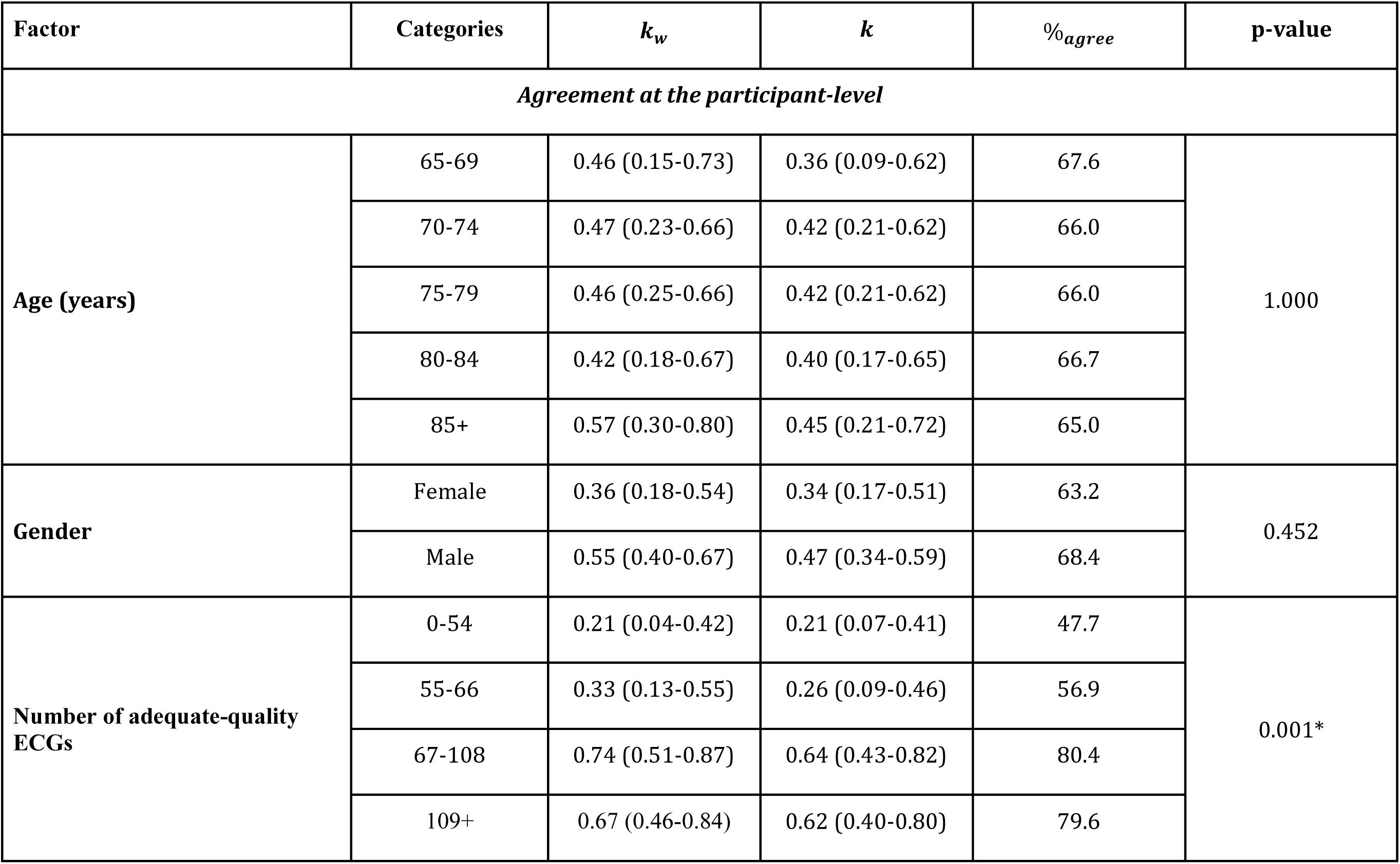
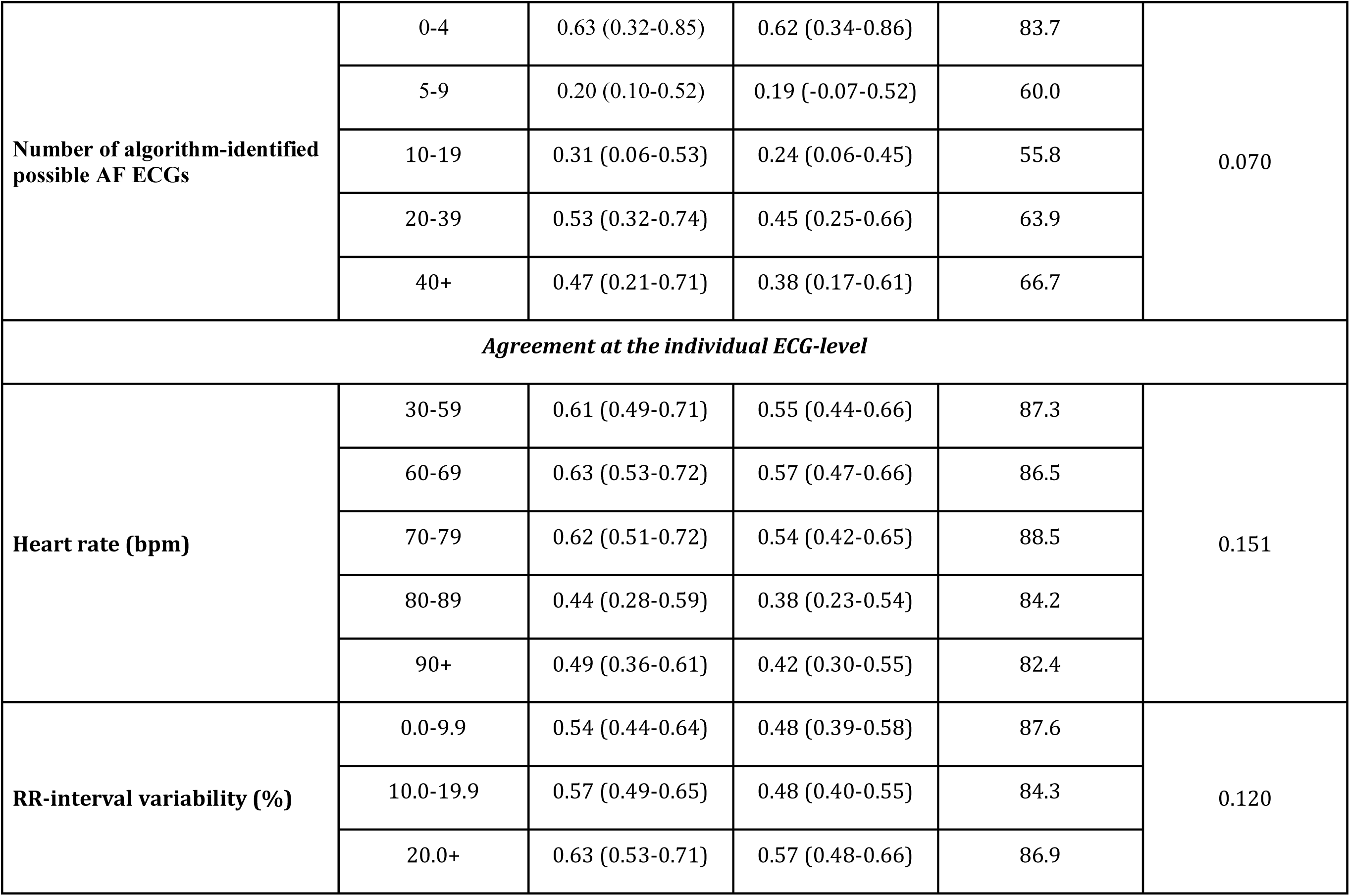
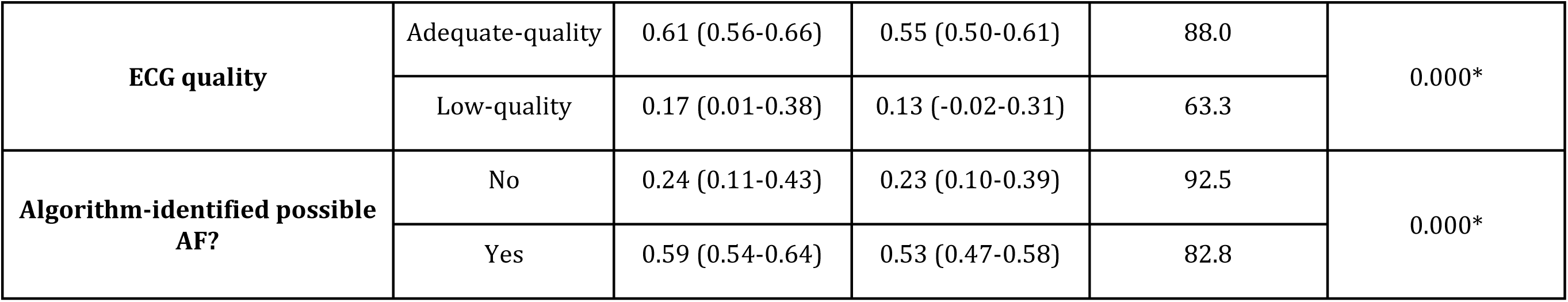
Relationships between the level of agreement between cardiologists and factors at the participant and ECG-level.

| Factor | Categories | $k_w$ | $k$ | %*agree* | p-value |
| --- | --- | --- | --- | --- | --- |
| *Agreement at the participant-level* |  |  |  |  |  |
| **Age (years)** | 65-69 | 0.46 (0.15-0.73) | 0.36 (0.09-0.62) | 67.6 | 1.000 |
|  | 70-74 | 0.47 (0.23-0.66) | 0.42 (0.21-0.62) | 66.0 |  |
|  | 75-79 | 0.46 (0.25-0.66) | 0.42 (0.21-0.62) | 66.0 |  |
|  | 80-84 | 0.42 (0.18-0.67) | 0.40 (0.17-0.65) | 66.7 |  |
|  | 85+ | 0.57 (0.30-0.80) | 0.45 (0.21-0.72) | 65.0 |  |
| **Gender** | Female | 0.36 (0.18-0.54) | 0.34 (0.17-0.51) | 63.2 | 0.452 |
|  | Male | 0.55 (0.40-0.67) | 0.47 (0.34-0.59) | 68.4 |  |
| **Number of adequate-quality ECGs** | 0-54 | 0.21 (0.04-0.42) | 0.21 (0.07-0.41) | 47.7 | 0.001* |
|  | 55-66 | 0.33 (0.13-0.55) | 0.26 (0.09-0.46) | 56.9 |  |
|  | 67-108 | 0.74 (0.51-0.87) | 0.64 (0.43-0.82) | 80.4 |  |
|  | 109+ | 0.67 (0.46-0.84) | 0.62 (0.40-0.80) | 79.6 |  |
| Number of algorithm-identified possible AF ECGs | 0-4 | 0.63 (0.32-0.85) | 0.62 (0.34-0.86) | 83.7 | 0.070 |
|  | 5-9 | 0.20 (0.10-0.52) | 0.19 (-0.07-0.52) | 60.0 |  |
|  | 10-19 | 0.31 (0.06-0.53) | 0.24 (0.06-0.45) | 55.8 |  |
|  | 20-39 | 0.53 (0.32-0.74) | 0.45 (0.25-0.66) | 63.9 |  |
|  | 40+ | 0.47 (0.21-0.71) | 0.38 (0.17-0.61) | 66.7 |  |
| Agreement at the individual ECG-level |  |  |  |  |  |
| Heart rate (bpm) | 30-59 | 0.61 (0.49-0.71) | 0.55 (0.44-0.66) | 87.3 | 0.151 |
|  | 60-69 | 0.63 (0.53-0.72) | 0.57 (0.47-0.66) | 86.5 |  |
|  | 70-79 | 0.62 (0.51-0.72) | 0.54 (0.42-0.65) | 88.5 |  |
|  | 80-89 | 0.44 (0.28-0.59) | 0.38 (0.23-0.54) | 84.2 |  |
|  | 90+ | 0.49 (0.36-0.61) | 0.42 (0.30-0.55) | 82.4 |  |
| RR-interval variability (%) | 0.0-9.9 | 0.54 (0.44-0.64) | 0.48 (0.39-0.58) | 87.6 | 0.120 |
|  | 10.0-19.9 | 0.57 (0.49-0.65) | 0.48 (0.40-0.55) | 84.3 |  |
|  | 20.0+ | 0.63 (0.53-0.71) | 0.57 (0.48-0.66) | 86.9 |  |
| **ECG quality** | Adequate-quality | 0.61 (0.56-0.66) | 0.55 (0.50-0.61) | 88.0 | 0.000* |
|  | Low-quality | 0.17 (0.01-0.38) | 0.13 (-0.02-0.31) | 63.3 |  |
| **Algorithm-identified possible AF?** | No | 0.24 (0.11-0.43) | 0.23 (0.10-0.39) | 92.5 | 0.000* |
|  | Yes | 0.59 (0.54-0.64) | 0.53 (0.47-0.58) | 82.8 |  |

### 3.2. Reliability of ECG interpretation

The inter-rater reliability of AF diagnosis at the individual ECG-level was moderate (*κ_w_* = 0.58 (0.53 − 0.63); *κ* = 0.51 (0.46 − 0.56); and %*_agree_* = 86.1% (Table 3). Referring to the ECG-level results in Figure 3 and Table 2, the level of agreement was significantly associated with ECG quality, with low-quality ECGs associated with a lower level of agreement. This remained regardless of whether quality was assessed using cardiologist comments on ECG quality (for which 94 ECGs, 5.1%, were deemed low-quality), the automated algorithm assessment (67 ECGs, 3.6%), or a combination of both (139 ECGs, 7.5%). The level of agreement was also significantly associated with whether or not an ECG exhibited algorithm-identified possible AF, where ECGs exhibiting possible AF were associated with a higher level of agreement. The was no significant association between the level of agreement and heart rate or RR-interval variability.

**Table 3:** Agreement between cardiologists on ECG-level AF diagnoses.

|  |  | Cardiologist 2 |  |  |  |
| --- | --- | --- | --- | --- | --- |
|  |  | AF | non-AF | cannot exclude AF |  |
| Cardiologist 1 | AF | 144 | 84 | 7 | 235 |
|  | non-AF | 28 | 1,424 | 37 | 1,489 |
|  | cannot exclude AF | 7 | 93 | 19 | 119 |
|  |  | 179 | 1,601 | 63 | 1,843 |

### 3.3. Comparison of cardiologists’ reviewing practices

The two cardiologists’ reviewing practices differed. At the participant-level, one cardiologist diagnosed more participants with AF than the other (72 out of 190, *i.e.* 38%, vs. 50, *i.e.* 26%) (see Table 1). Similarly, at the ECG-level, this cardiologist diagnosed more ECGs as AF than the other (235 out of 1,843, *i.e.* 13%, vs. 179, *i.e.* 9.7%), and more ECGs as ‘cannot exclude AF’ than the other (119, *i.e.* 6%, vs. 63, *i.e.* 3%) (see Table 3). Most of the ECGs diagnosed as AF by the cardiologists exhibited an irregular rhythm as identified by the algorithm (95% of the 235 ECGs diagnosed as AF by one cardiologist, 88% of the 179 ECGs diagnosed as AF by the other cardiologist, and 95% of the 137 ECGs diagnosed as AF by both cardiologists). Examples of ECGs are provided in Figures 4 and 5: Figure 4 shows ECGs on which there was complete agreement of (a) non-AF and (b) AF; Figure 5 shows ECGs on which there was complete disagreement with one cardiologist diagnosing AF and the other non-AF.

**Figure 4:**
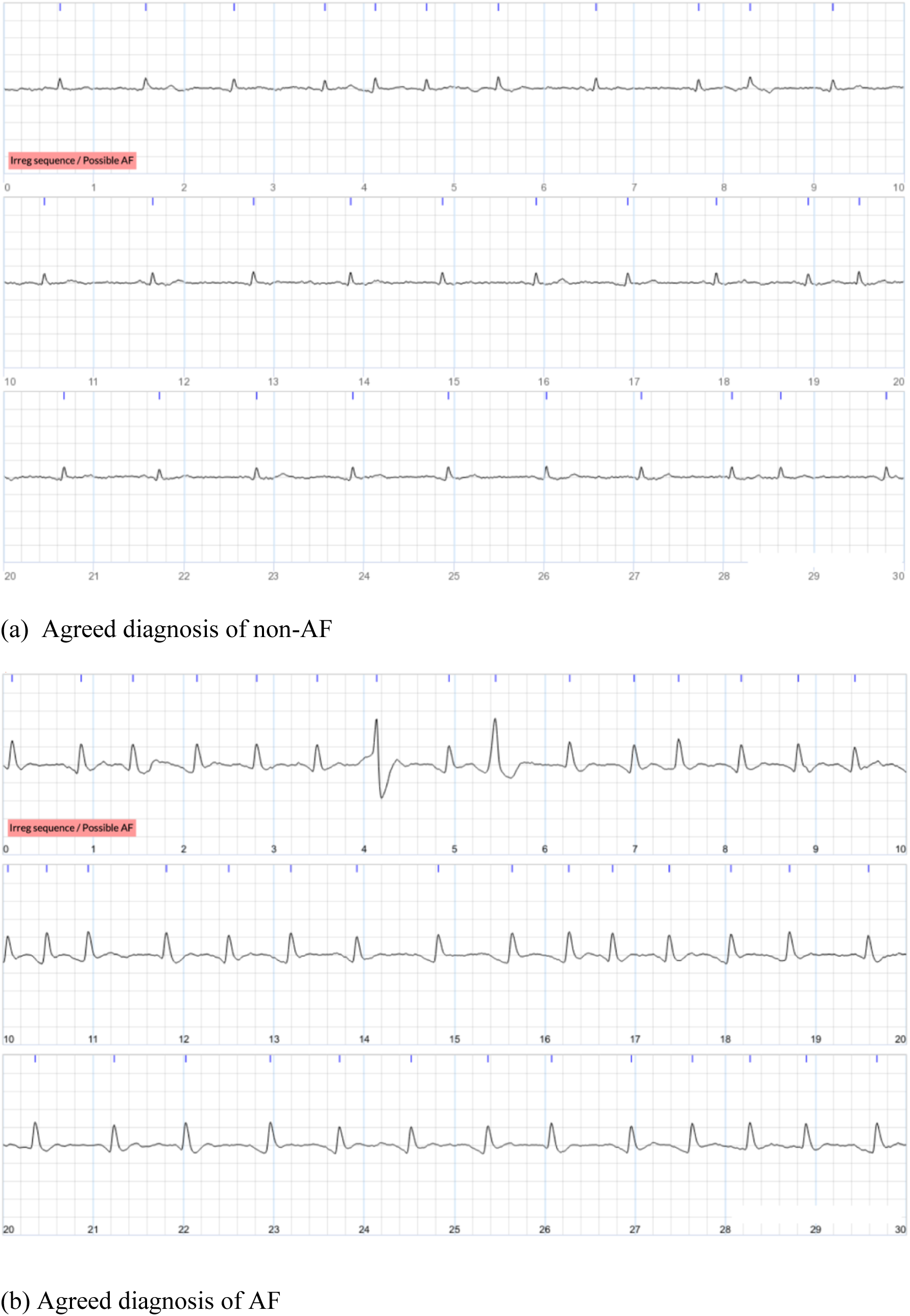
Examples of single-lead ECGs on which the cardiologists agreed. Each image shows a 30-second ECG, with 10 seconds per line.

**Figure 5:**
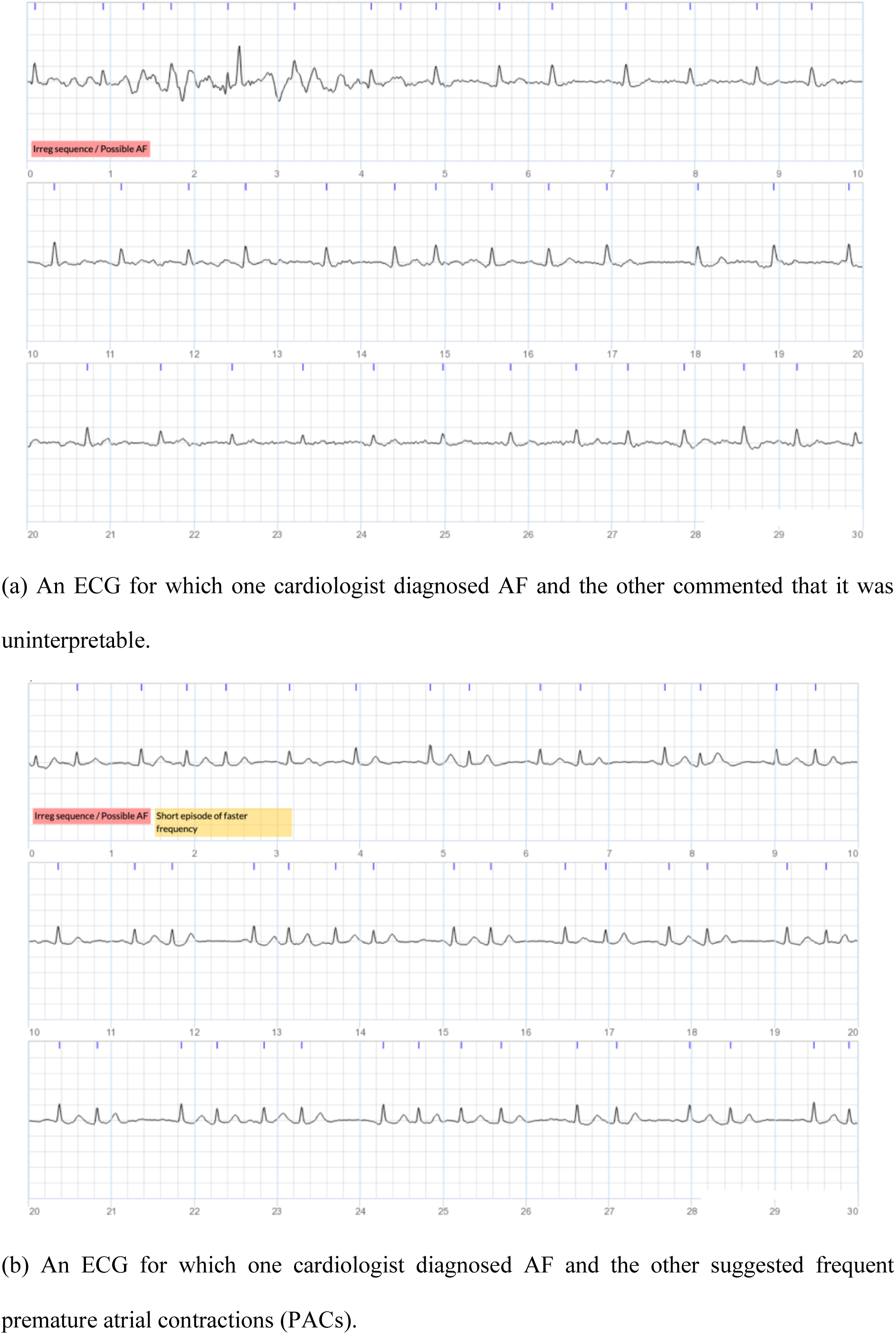
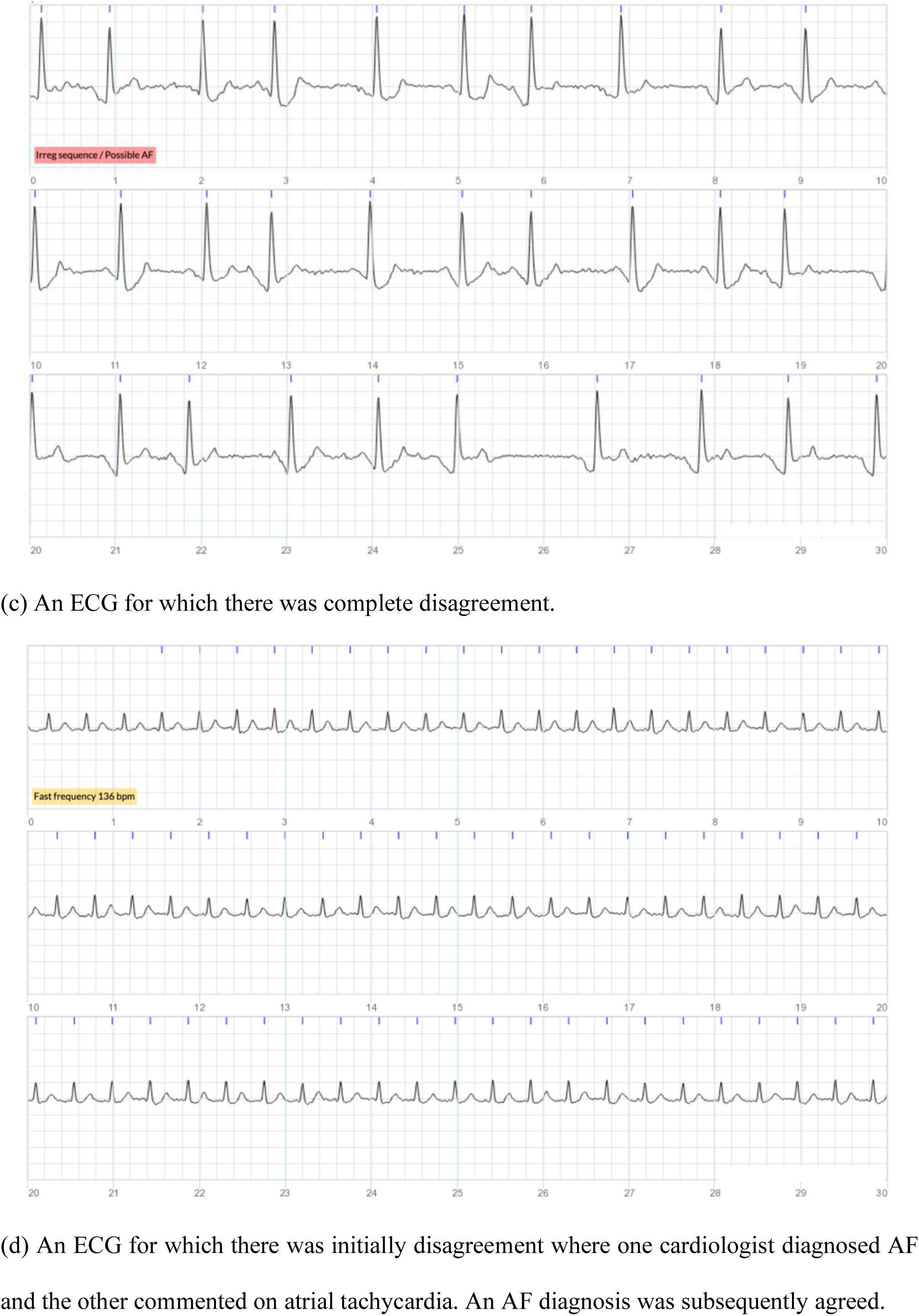
Examples of single-lead ECGs over which the cardiologists completely disagreed, with Cardiologist 1 diagnosing AF, and Cardiologist 2 diagnosing non-AF. Each image shows a 30-second ECG, with 10 seconds per line.

## 4. Discussion

### 4.1. Summary of findings

This study provides evidence on the inter-rater reliability of single-lead electrocardiogram interpretation, and the factors that influence this. Moderate agreement was observed between cardiologists on participant-level diagnoses of AF in a population-based AF screening study when this diagnosis was made using multiple ECGs per participant. The key factor associated with the level of agreement at the participant-level was the number of adequate-quality ECGs recorded by a participant, with higher levels of agreement in those who recorded more adequate-quality ECGs. Moderate agreement was observed between cardiologists on the diagnoses of individual ECGs. Similarly, at the ECG-level, low-quality ECGs were associated with lower levels of agreement. In addition, lower levels of agreement were observed on those ECGs not exhibiting algorithm-identified possible AF.

### 4.2. Comparison with existing literature

The levels of agreement in AF diagnosis from single-lead ECGs observed in this study are lower than in many previous studies. Previous studies have found almost perfect agreement when interpreting 12-lead ECGs, but lower levels of agreement when interpreting single-lead ECGs. In an analysis of 12-lead ECGs from the SAFE AF Screening Trial, cardiologists agreed on the diagnosis of 99.7% of ECGs (all but 7 of 2,592 analysed ECGs) (16). In comparison, in the present study of single-lead ECGs cardiologists agreed on the diagnosis of 86.1% of ECGs (1,587 out of 1,843 ECGs). However, the proportion of normal ECGs included in this study was substantially lower than in the SAFE AF Screening Trial (less than 1% in this study, versus 93% in SAFE), so the simple level of agreement is not directly comparable. Similarly, in a study of the diagnosis of supraventricular tachycardia in hospital patients, an almost perfect agreement of *κ* = 0.97 was observed in interpretation of 12-lead ECGs, compared to a substantial agreement of *κ* = 0.76 when using single-lead ECGs from the same patients (17). The previously reported levels of agreement for the diagnosis of AF from single-lead ECGs have varied greatly between studies: fair agreement was observed by Kearley *et al.* (18) (*κ* = 0.28); moderate agreement was observed by Lowres *et al.* (19) (weighted *κ* = 0.4); substantial agreements were observed by Poulsen *et al.* (12) (*κ* = 0.65) and Kearley *et al.* (18) (*κ* = 0.76); and almost perfect agreements were observed by *Desteghe et al.* (11)(*κ* = 0.69 to 0.86), *Koshy et al.* (20) (*κ* = 0.80 to 0.83), Wegner *et al.* (13) (*κ* = 0.90), and Racine *et al.* (21)(*κ* = 0.94). The variation in levels of agreement may have been contributed to by study setting and underlying frequency of AF, since those studies which reported the lowest levels of agreement took place out-of-hospital (18, 19). The present study, conducted in the community, similarly observed lower levels of agreement than many other studies (*κ* = 0.42 at the participant-level, and *κ* = 0.51 at the ECG-level). In the context of AF screening, a 69.2% level of agreement has been reported in a previous AF screening study by Pipilas *et al.* (22), compared to 86.1% in the present screening study. The low levels of agreement in the present study could have been contributed to by: (i) the ECGs being more challenging to review as an algorithm and a nurse filtered out most ECGs which did not exhibit signs of AF (and are therefore easier to interpret) prior to cardiologist review; (ii) the ECGs being of lower quality since participants recorded ECGs themselves without clinical supervision; and (iii) the use of an additional diagnostic category of ‘cannot exclude AF’.

This study’s findings about factors which influence the reliability of ECG interpretation complement those reported previously (11,12,13,22,23). It has previously been reported that ECGs exhibiting baseline wander, noise, premature beats, or low-amplitude atrial activity are associated with mis-diagnoses (11,23). In this study low-quality ECGs were similarly associated with lower levels of agreement between cardiologists. The significant proportion of low-quality ECGs obtained when using a handheld ECG device has been reported previously, with 12% of ECGs being judged as ‘very low quality’ in (22), 13% as ‘not useable’ in (12), and 20% as ‘inadequate quality’ in (13). In comparison, in this study 7.5% of ECGs were deemed low-quality according to the cardiologists or algorithm, which likely represents an underestimate since cardiologist labels were obtained on an ad-hoc basis. In contrast, in the STROKESTOP study, which also utilised the Zenicor One device, only 0.99% of ECGs were classified by the algorithm as low quality (9), as compared to 3.6% in our study. Differences in population (SAFER: people aged 70 and over with no upper age limit; STROKESTOP: people aged 75 or 76 years) or training (SAFER: in general practice; STROKESTOP: in a screening centre) may have contributed to this difference.

The accuracy of both automated and manual diagnosis of AF from single-lead ECGs has been assessed previously. A recent meta-analysis found pooled sensitivities and specificities of automated ECG diagnoses of 89% and 99% respectively in the community setting (24). The accuracy of manual diagnoses has varied greatly between previous studies, with sensitivities and specificities in comparison to reference 12-lead ECGs reported as: 77.4% and 73.0% (22), 90% and 79% (21), 76-92% and 84-100% (20), 89-100% and 85-88% (25), 92.5% and 89.8% (26), 93.9% and 90.1% (18), 100% and 94% (13), and 100% and 100% (27). In all of these studies, the single-lead ECGs were recorded under supervision. In contrast, the present study considered ECGs collected using a telehealth device at home without supervision.

### 4.3. Strengths and limitations

There are several strengths to this study. First, we assessed the level of agreement in both participant-level ECG-level AF diagnoses which is of particular relevance in AF screening, whereas most previous work has been limited to ECG-level diagnoses. Second, the ECGs used in this study were collected in a prospective population-based AF screening study, and are therefore representative of ECGs captured in telehealth settings by older adults without clinical supervision. The ECGs were recorded using dry electrodes, as opposed to the gel electrodes used in clinical settings. Dry electrodes can result in poorer conduction and therefore lower signal quality, making interpretation more challenging. Since smartwatches also use dry electrodes, the findings are expected to be relevant to the growing use of ECG-enabled consumer devices. Third, the ECGs included in the analysis are representative of those which would be sent for clinical review in real-world settings: ECGs without signs of abnormalities were excluded using an automated, CE-marked analysis system, leaving only those ECGs with signs of abnormalities for review. Fourth, the study included a large number of ECGs (1,843), each interpreted by two cardiologists. Fifth, we used Cohen’s kappa statistic to assess the level of agreement between cardiologists: this statistic takes into account agreement by chance unlike the percentage agreement (Viera and Garrett, 2005).

The key limitations to this study are as follows. First, the findings are based on data from only 190 participants who had an abnormal ECG flagged in the study, and mostly abnormal ECGs (*i.e.* the less straightforward to review). Second, inter-rater agreement was assessed using diagnoses provided by only two cardiologists. Whilst the findings would be more generalisable with additional cardiologists, we anticipate that the observed levels of agreement are towards the higher end of the range of expected levels of agreement since the cardiologists in this study were highly experienced. Indeed, both had considerable prior experience of reviewing ECGs from handheld devices, although no formal comparison of their experience was made prior to the study. Indeed, the percentage agreement at the ECG-level in this study (86.1%) was higher than the analogous mean variability reported in an analysis of 15 cardiologists reviewing VITAL-AF data (22) where pairs of cardiologists agreed on 69% of ECGs. Third, not all ECGs sent for review were interpreted by both cardiologists, with those not interpreted by both cardiologists excluded from the analysis. Fourth, the initial diagnosis was not recorded for a small minority of the ECGs reviewed by both cardiologists (29 out of 1,872, 1.5%), so these were not included in the analysis. Fifth, the investigation of factors influencing the level of agreement was limited to those factors collected during a screening programme, and the study may have been underpowered to identify some further associations: future research may elucidate further factors which were not identified in this study. Sixth, we did not investigate intra-rater variability in this study, which may change over time as more experience is gained. Seventh, this study was conducted using ECGs recorded using one particular device after in-person training, and it is not clear how generalisable the findings are to other devices or with other levels of training, which may impact ECG quality. Finally, it should be remembered that the study assessed the reliability of ECG interpretation (*i.e.* the level of agreement between two cardiologists), rather than the accuracy of ECG interpretation (*i.e.* a comparison of cardiologist interpretation against an independent reference). In doing so, the study identified factors associated with reduced levels of agreement, providing evidence on how to improve the level of agreement, and subsequently the reliability of interpretation.

### 4.4. Implications

This study indicates that steps should be taken to ensure diagnoses based on single-lead ECGs are as reliable as possible. Out of 2,141 participants screened for AF, there was agreement between cardiologists on diagnoses of AF for 44 participants, complete disagreement for 31 participants (AF vs. non-AF), and partial disagreement for 33 participants (AF or non-AF vs. cannot exclude AF). In terms of disease prevalence, there was agreement on AF diagnosis in 2.1% of the sample population, complete disagreement in 1.4%, and partial disagreement in 1.5%.

The findings could inform the design of AF screening programmes. AF screening programmes often include collection of multiple short ECGs (or a continuous ECG recording) over a prolonged period to capture even infrequent episodes of paroxysmal AF. The results of this study indicate that a prolonged period is also required to obtain reliable diagnoses: at least 67 adequate-quality ECGs were required for a reliable diagnosis in this study, providing evidence that screening programmes should be designed to capture at least this many adequate-quality ECGs from all participants (*i.e.* at least 17 days of screening when recording 4 ECGs per day, and potentially 21 days of screening to account for missed or low-quality ECGs). In addition, no association was found between participant gender or age and the reliability of diagnoses, indicating that it is reasonable to use single-lead ECGs in older adults of a wide range of ages (from 65 to 90+ in this study).

The findings of this study could also underpin strategies to obtain more reliable participant-level diagnoses through personalised screening. Those individuals who are likely to receive a less reliable diagnosis could be identified by using an automated algorithm to analyse the quality of incoming ECGs, and then the duration of screening could be extended in those individuals without sufficient adequate-quality ECGs. This could help increase reliability by increasing the number of adequate-quality ECGs available for diagnosis. Second, participants with a high proportion of low-quality ECGs could be offered additional training on ECG measurement technique, potentially by telephone.

This study highlights the need to ensure single-lead ECG interpreters receive sufficient training. The ECGs in this study were interpreted by highly experienced cardiologists, and yet there was still disagreement over diagnoses for 16% of those participants sent for cardiologist review. If single-lead ECG-based AF screening is widely adopted in the future, then it will be important to ensure all ECG interpreters receive sufficient training and gain sufficient experience in single-lead ECG interpretation to provide reliable diagnoses. We note that single-lead ECG interpretation presents additional challenges beyond those encountered in 12-lead ECG interpretation: ECGs may be of lower quality (12), P-waves may not be as visible (11), and only one lead is available. It is notable that in a hospital-conducted study where 12-lead ECGs were performed at the same time as Zenicor One ECGs, independent reading of the single lead ECGs by two senior cardiologists with a review by a third where there was disagreement, resulted in a sensitivity of 98% and specificity of 99% for AF present on the 12 lead ECG (10). This suggests that accurate (and therefore reliable) single lead ECG interpretation is possible, raising the prospect that training might be effective.

The findings of this study indicate that it is important that the quality of single-lead ECGs is as high as possible. Particularly given the implications of an AF diagnosis such as recommendations for anticoagulation treatment which increases the risk of bleeding. The development of consumer and telehealth ECG devices involves making a range of design decisions which can influence the quality of ECGs sent for clinical review, including: the size, type, and anatomical position of electrodes; the filtering applied to signals to reduce noise; and whether to exclude ECGs of insufficient quality from clinical review (and if so, how best to identify these ECGs). Device should designers consider the potential effects of these design decisions on the reliability of diagnoses.

## 5. Conclusion

Moderate agreement was found between cardiologists when diagnosing AF from single-lead ECGs in an AF screening study. The study indicates that for every 100 screening participants diagnosed with AF by two cardiologists, there would be complete disagreement over the diagnosis of 70 further participants. This provides great incentive for ensuring that the interpretation of single-lead ECGs is as reliable as possible. Key factors were identified which influence the reliability of single-lead ECG interpretation. Most importantly, the quality of ECG signals greatly influenced reliability. In addition, when multiple ECGs were acquired from an individual, the reliability of participant-level diagnoses was influenced by the number of adequate-quality ECGs available for interpretation. This new evidence could help improve single-lead ECG interpretation, and consequently increase the effectiveness of screening for AF using single-lead ECG devices. Future work should investigate how to obtain ECGs of the highest possible quality in the telehealth setting, and how best to train ECG interpreters to ensure diagnoses are as accurate as possible.

## Data Availability

Requests for pseudonymised data should be directed to the SAFER study co-ordinator (Andrew Dymond using) and will be considered by the investigators, in accordance with participant consent.

## Acknowledgment

This study is funded by the National Institute for Health and Care Research (NIHR), Programme Grants for Applied Research Programme (Reference Number RP-PG0217-20007); the NIHR School for Primary Care Research (SPCR-2014-10043, project 410); and the British Heart Foundation (BHF) grant number FS/20/20/34626. The views expressed are those of the authors and not necessarily those of the NIHR or the Department of Health and Social Care.

## References

1. Wilson FN, Kossmann CE, Burch GE, Goldberger E, Graybiel A, Hecht H, et al. Recommendations for Standardization of Electrocardiographic and Vectorcardiographic Leads. Circulation. 1954;10(4):564–573.

2. Rafie N, Kashou AH, Noseworthy PA. ECG Interpretation: Clinical Relevance, Challenges, and Advances. Hearts. 2021;2(4):505–513.

3. Hall A, Mitchell ARJ, Wood L, Holland C. Effectiveness of a single lead AliveCor electrocardiogram application for the screening of atrial fibrillation. Medicine (Baltimore). 2020;99(30):e21388.

4. Wolf PA, Abbot RD, Kannel WB. Atrial fibrillation as an independent risk facor for stroke: the Framingham study. Stroke. 1991;22:983–8.

5. Ruff CT, Giugliano RP, Braunwald E, Hoffman E, Deenadayalu N, Ezekowitz M, et al. Comparison of the efficacy and safety of new oral anticoagulants with warfarin in patients with atrial fibrillation: a meta-analysis of randomised trials. The Lancet. 2014;383(9921):955–962.

6. Public Health England. Atrial fibrillation prevalence estimates in England: Application of recent population estimates of AF in Sweden. PHE Publications Gateway Number: 2014778. PHE Publications Gateway Number 2014778; 2015.

7. Svennberg E, Engdahl J, Al-Khalili F, Friberg L, Frykman V, Rosenqvist M. Mass screening for untreated atrial fibrillation: the STROKESTOP study. Circulation. 2015;131(25):2176–84.

8. Hindricks G, Potpara T, Dagres N, Bax JJ, Boriani G, Dan GA, et al. 2020 ESC Guidelines for the diagnosis and management of atrial fibrillation developed in collaboration with the European Association for Cardio-Thoracic Surgery (EACTS). Eur Heart J. 2021;42(5):373–498.

9. Svennberg E, Stridh M, Engdahl J, Al-Khalili F, Friberg L, Frykman V, et al. Safe automatic one-lead electrocardiogram analysis in screening for atrial fibrillation. Europace. 2017;19(9):1449–53.

10. Malmqvist J, Engdahl J, Sjölund G, Doliwa P. Sensitivity and specificity of handheld one lead ECG detecting atrial fibrillation in an outpatient clinic setting. Journal of Electrocardiology 2024 83:106–110.

11. Desteghe L, Raymaekers Z, Lutin M, Vijgen J, Dilling-Boer D, Koopman P, et al. Performance of handheld electrocardiogram devices to detect atrial fibrillation in a cardiology and geriatric ward setting. EP Eur. 2017 Jan 1;19(1):29–39.

12. Poulsen MB, Binici Z, Dominguez H, Soja AMB, Kruuse C, Hornnes AH, et al. Performance of short ECG recordings twice daily to detect paroxysmal atrial fibrillation in stroke and transient ischemic attack patients. Int J Stroke. 2017;12(2):192–6.

13. Wegner FK, Kochhäuser S, Ellermann C, Lange PS, Frommeyer G, Leitz P, et al. Prospective blinded Evaluation of the smartphone-based AliveCor Kardia ECG monitor for Atrial Fibrillation detection: The PEAK-AF study. Eur J Intern Med. 2020 Mar 1;73:72–5.

14. Pandiaraja M, Brimicombe J, Cowie M, Dymond A, Lindén HC, Lip GYH, et al. Screening for atrial fibrillation: Improving efficiency of manual review of handheld electrocardiograms. Eng Proc. 2020;2(1):78.

15. Viera AJ, Garrett JM. Understanding interobserver agreement: the kappa statistic. Fam Med. 2005 May;37(5):360–3.

16. Mant J, Fitzmaurice DA, Hobbs FDR, Jowett S, Murray ET, Holder R, et al. Accuracy of diagnosing atrial fibrillation on electrocardiogram by primary care practitioners and interpretative diagnostic software: Analysis of data from screening for atrial fibrillation in the elderly (SAFE) trial. Br Med J. 2007;335(7616):380–2.

17. Wegner FK, Kochhäuser S, Frommeyer G, Lange PS, Ellermann C, Leitz P, et al. Prospective blinded evaluation of smartphone-based ECG for differentiation of supraventricular tachycardia from inappropriate sinus tachycardia. Clinical Research in Cardiology. 2021;110(6):905–12.

18. Kearley K, Selwood M, Bruel AV den, Thompson M, Mant D, Hobbs FR, et al. Triage tests for identifying atrial fibrillation in primary care: a diagnostic accuracy study comparing single-lead ECG and modified BP monitors. BMJ Open. 2014 Apr 1;4(5):e004565.

19. Lowres N, Neubeck L, Salkeld G, Krass I, McLachlan AJ, Redfern J, et al. Feasibility and cost-effectiveness of stroke prevention through community screening for atrial fibrillation using iPhone ECG in pharmacies: The SEARCH-AF study. Thrombosis and Haemostasis. 2014;111(6):1167–76.

20. Koshy AN, Sajeev JK, Negishi K, Wong MC, Pham CB, Cooray SP, et al. Accuracy of blinded clinician interpretation of single-lead smartphone electrocardiograms and a proposed clinical workflow. American Heart Journal. 2018;205:149–53.

21. Racine HP, Strik M, Zande J van der, Alrub SA, Caillol T, Haïssaguerre M, et al. Role of Coexisting ECG Anomalies in the Accuracy of Smartwatch ECG Detection of Atrial Fibrillation. Canadian Journal of Cardiology. 2022 Nov 1;38(11):1709–12.

22. Pipilas DC, Khurshid S, Atlas SJ, Ashburner JM, Lipsanopoulos AT, Borowsky LH, et al. Accuracy and variability of cardiologist interpretation of single lead electrocardiograms for atrial fibrillation: The VITAL-AF trial. American Heart Journal. 2023 Nov 1;265:92–103.

23. Davidenko JM, Snyder LS. Causes of errors in the electrocardiographic diagnosis of atrial fibrillation by physicians. J Electrocardiol. 2007 Sep 1;40(5):450–6.

24. Wong KC, Klimis H, Lowres N, Huben A von, Marschner S, Chow CK. Diagnostic accuracy of handheld electrocardiogram devices in detecting atrial fibrillation in adults in community versus hospital settings: a systematic review and meta-analysis. Heart. 2020 Aug 1;106(16):1211–7.

25. Ford C, Xie CX, Low A, Rajakariar K, Koshy AN, Sajeev JK, et al. Comparison of 2 Smart Watch Algorithms for Detection of Atrial Fibrillation and the Benefit of Clinician Interpretation: SMART WARS Study. JACC: Clinical Electrophysiology. 2022 Jun 1;8(6):782–91.

26. Karregat EPM, Himmelreich JCL, Lucassen WAM, Busschers WB, van Weert HCPM, Harskamp RE. Evaluation of general practitioners’ single-lead electrocardiogram interpretation skills: a case-vignette study. Family practice. 2021;38(2):70–5.

27. Himmelreich JCL, Karregat EPM, Lucassen WAM, Weert HCPM van, Groot JR de, Handoko ML, et al. Diagnostic Accuracy of a Smartphone-Operated, Single-Lead Electrocardiography Device for Detection of Rhythm and Conduction Abnormalities in Primary Care. The Annals of Family Medicine. 2019 Sep 1;17(5):403–11.

28. Mant J, Modi R, Charlton P, Dymond A, Massou E, Brimicombe J, et al. The feasibility of population screening for paroxysmal atrial fibrillation using handheld ECGs. EP Europace. 2024;26(3):euae056.

29. Lopez Perales CR, Van Spall HGC, Maeda S, Jimenez A, Laţcu DG, Milman A, et al. Mobile health applications for the detection of atrial fibrillation: A systematic review. EP Europace. 2021;23(1):11–28.

